# Toward a more comprehensive understanding of network centrality disruption in amnestic Mild Cognitive Impairment: a MEG multilayer approach

**DOI:** 10.1101/2024.01.30.24302028

**Authors:** Ignacio Taguas, Sandra Doval, Fernando Maestú, David López-Sanz

## Abstract

Alzheimer’s Disease (AD) is the most common form of dementia. Its early stage, amnestic Mild Cognitive Impairment (aMCI), is characterized by disrupted information flow in the brain. Previous studies using electrophysiological techniques to investigate AD’s functional connectivity changes have yielded inconsistent results; contributing factors may include the specific metric employed and the separate study of brain activity in each frequency band. Our study addresses this by employing a cross-frequency approach to compare functional networks in 172 healthy subjects and 105 aMCI patients. Using magnetoencephalography, we constructed source-based multilayer graphs considering both intra- and inter-band functional connectivity across the five classical frequency bands. We assessed network changes through three centrality measures (strength, eigenvector centrality, and betweenness centrality), and combined them into a unified centrality score for a comprehensive assessment of centrality disruption in aMCI. Results revealed a notable shift in centrality distribution in aMCI patients spatially and across frequencies. Posterior brain regions decrease synchrony between high-frequency oscillations and other regions’ activity across all frequencies, while anterior regions increase synchrony between low-frequency oscillations and other regions’ activity across all bands. Thus, posterior regions reduce their relative importance in favor of anterior regions. This study demonstrates that considering the interplays between different frequency bands enhances understanding of AD network dynamics and sets a precedent for multilayer functional network studies. Our findings provide valuable insights into the intricate changes that occur in functional brain networks during the early stages of AD, offering a framework for developing interventions aimed at modifying the disease trajectory.

**Significance statement:** Over 55 million people worldwide suffer from Alzheimer’s Disease. The brain changes causing the symptoms begin around 20 years before their onset, so characterizing these changes in the early stage, mild cognitive impairment, is of utmost importance. Magnetoencephalography and electroencephalography (M/EEG) are two commonly used techniques to assess changes in functional networks; however, the existing studies yield inconsistent results. Our study applies a novel methodology for the analysis of M/EEG data that addresses two factors contributing to this effect: the metric employed to assess network changes and the use of a cross-frequency approach. Results show that posterior regions reduce their relative importance in favor of anterior regions. This study sets a precedent for the functional study of all brain disorders.

## 1 Introduction

Alzheimer’s Disease (AD) is an incurable neurodegenerative disease that accounts for about two-thirds of dementia cases (Duong et al., 2017). The clinical manifestations start with a progressive loss of episodic memory, followed by impairment of other cognitive domains (Silva et al., 2019). But clinical symptoms are only the tip of the iceberg, as they appear ten to fifteen years after the earliest neuronal dysfunctions occur (Khanahmadi et al., 2015). The cascade of events leading to AD starts with the malfunctioning of two key proteins, amyloid-β (Aβ) and tau, which accumulate to form extracellular Aβ plaques and intracellular tau neurofibrillary tangles. These aggregates affect synapses, altering neuronal communication, and as more synapses disappear, functional networks are disrupted, ultimately causing cognitive dysfunctions. Thus, brain network disruption constitutes a potential marker for the detection of AD even before clinical symptoms manifest, and its study in AD and its prodromal state, Mild Cognitive Impairment (MCI), is crucial.

Brain networks are commonly studied through functional connectivity: the statistical dependence through time between physiological signals from different brain regions (Friston, 2011). Functional connectivity is a direct measure of brain synchrony. Increments in functional connectivity reflect an increase in brain synchrony, while decreases imply the opposite; both changes can be pathological.

Functional connectivity disruption in AD and MCI patients is a common finding. However, previous literature shows erratic results due to conflicting methodological factors (including differences in the various functional connectivity metrics, inconsistencies in diagnostic criteria, and inadequacies in sample sizes) (Briels et al., 2020). Here, we review the most persistent results. The anterior aspects of the brain exhibit hypersynchrony in patients in delta and theta frequencies (Ruiz-Gómez et al., 2019; Ranasinghe et al., 2020), and sometimes in alpha (López-Sanz et al., 2017). The posterior aspects of the brain exhibit hyposynchrony in patients in alpha and beta (Núnez et al., 2019; Smailovic et al., 2020), and seldom in gamma (Cuesta et al., 2022). In the earliest (preclinical) stages of the disease, studies report hypersynchrony in theta and delta in posterior brain regions (Nakamura et al., 2017).

A further step in the study of functional networks is modeling them as graphs. A graph is a set of elements, called nodes, connected by links. Under this framework, nodes denote brain regions and links denote functional connectivity values. One of the main advantages of graphs is that they allow us to study the importance of each neural substrate in the network by measuring their centrality. In graph theory, centrality is a vague concept that ranks nodes according to the role they play in the graph; different centrality measures use different criteria.

Analyses using graph theory imply a greater level of abstraction and methodological complexity than those directly measuring functional connectivity, which further complicates the interpretation of the results. Consequently, existing papers reach even more conflicting conclusions (Kucikova et al., 2021). Even so, it is a common finding that variations in cerebrospinal fluid biomarkers, namely Aβ and tau, entail centrality changes in MCI and AD (Binnewijzend et al., 2014; Ranasinghe et al., 2020).

All the articles mentioned in the previous paragraphs treat the different frequency bands individually. Separating brain signals into discrete and isolated frequency bands is convenient, as each band has been associated with specific physiological processes. Nonetheless, this strict division represents an oversimplification of brain dynamics. Several authors suggest that studying frequency-specific functional graphs in isolation might play a role in the dissimilar results between studies and hint at the use of a more integrative approach: the use of multilayer graphs, in which each layer corresponds to a frequency band. These graphs consider the interplays between the different frequency bands, as they contain both intra- and inter-band connections. In a recent study, Yu et al. (2017) concluded that MEG-based brain graphs comprising all bands reveal information that cannot be extracted by studying the frequency graphs separately, although this work solely focused on intra-band connections.

Previous studies have applied this approach to the study of disorders such as AD (Yu et al., 2017) and schizophrenia (De Domenico et al., 2016), but not MCI. In the present work, we have approached centrality changes, measured by several widely used centrality metrics, in amnestic MCI patients, considering functional connectivity interactions between all pairs of nodes in all frequency bands. To prevent oversimplifying the network structure, we constructed the graphs using brain sources as nodes and the original functional connectivity values as links, without applying any transformations (such as thresholding or binarization). We expect MCI participants to exhibit centrality disruptions affecting their whole brain communication patterns. Since previous studies suggest non-linear trajectories during AD, we anticipate both centrality increases and decreases depending on the brain region.

## 2 METHODS

### 2.1 Participants

The study participants were recruited from three different locations in Madrid, Spain: Hospital Clínico San Carlos, Centro de Prevención del Deterioro Cognitivo, and Centro de Mayores del Distrito de Chamartin. The sample comprised 277 participants: 172 healthy controls and 105 MCI patients. Table 1 summarizes the most important demographic and neuropsychological characteristics.

**TABLE 1.** Descriptive characteristics of the two groups.

| Variable | Controls | MCI |
| --- | --- | --- |
| Age (years) | 69.3 ± 5.3 | 74 ± 4.6* |
| Sex (F – M) | 105 – 67 | 69 – 36 |
| Education (years) | 14.6 ± 5.2 | 8.5 ± 4.7* |
| MMSE | 29.2 ± 1 | 26.5 ± 2.5* |
| Inverse digits | 6.2 ± 2.1 | 4 ± 1.4* |
| BNT | 54.2 ± 6.2 | 44.7 ± 10.5* |
| TMT-B (seconds) | 104.9 ± 57.4 | 233.7 ± 103.3* |
| LM-immediate | 39.3 ± 11.9 | 15.4 ± 7.9* |
| LM-delayed | 23.8 ± 9.3 | 6 ± 5.9* |

\*P<.001 (P (sex) > 0.05); Mann-Whitney non-parametric test or Pearson's $\chi^2$ test, as appropriate.
Data presented as mean ± SD, except for sex.
Abbreviations: MCI, Mild Cognitive Impairment; MMSE, Mini Mental State Examination; BNT, Boston Naming Test; TMT-B, Trail Making Test B; LM, Logical memory.

To assess the general cognitive and functional status of each participant, a group of experts consisting of neuropsychologists, psychiatrists, and neurologists administered a set of screening questionnaires: Mini Mental State Examination (MMSE), Geriatric Depression Scale – Short Form (GDS-SF), Functional Assessment Questionnaire and Hachinski Ischemic Score. They then interviewed each participant and performed a thorough neuropsychological assessment. Based on this information, the evaluators placed each participant in a specific diagnostic group; participants were diagnosed with MCI using the criteria outlined by Petersen and Grundman (Grundman et al., 2004). Also, only those MCI patients who had memory impairment were included in the study.

The sample for the analyses only included participants aged 60 years or older. The exclusion criteria employed were the following: (1) history of psychiatric or neurological disorders, (2) evidence of infection, infarction or focal lesions in a T2-weighted scan within 2 months prior to the MEG acquisition, (3) consumption of drugs that could affect MEG recordings (e.g., cholinesterase inhibitors), (4) alcoholism, (5) chronic use of anxiolytics, narcotics, anticonvulsants or sedative hypnotics, (6) a modified Hachinski score of 5 or more, and (7) a GDS-SF score of 5 or higher. Additional analyzes were carried out to discard causes of MCI other than dementia (diabetes mellitus, thyroid problems, vitamin B12 deficit, syphilis, and HIV). For the control group, participants with an MMSE score of less than 27 were excluded, as well as those who reported Subjective Memory Complaints in the clinical interview.

Informed consent was obtained from all participants. The ethics committee of Hospital Clínico San Carlos approved the study, and the procedure was carried out in accordance with the approved guidelines and regulations.

### 2.2 MEG recordings

We recorded brain activity using a 306 channel Vectorview MEG system (Elekta AB, Stockholm, Sweden), which consisted of 102 magnetometers and 204 planar gradiometers. The MEG system was housed in a field-isolated room (VacuumSchmelze GmmbH, Hanau, Germany) at the Center for Biomedical Technology in Madrid, Spain.

Recordings were obtained under eyes-closed resting-state conditions and lasted 4 to 5 minutes, at a 1000 Hz sampling rate with an online anti-alias bandpass filter (cut-off frequency filtering of 0.1 to 330 Hz). We used a 3D Fastrak Polhemus digitizer to computerize the head shape, which captured around 300 points on the scalp surface and three anatomical reference points (nasion and left and right preauricular points). During the register, the device also recorded head position using four head position indication coils placed on the forehead and mastoids. Eye movements and blinks were registered using two bipolar electrooculogram electrodes placed above and below the left eye and a ground electrode attached to the upper cheek.

### 2.3 Preprocessing

We preprocessed the electrophysiological data using the Fieldtrip toolbox version 20170501 (Oostenveld et al., 2011) and in-house Matlab scripts (version 2021b). The pipeline included the following steps: (1) detecting and removing noisy channels using Maxfilter software, which virtually reconstructed their activity; (2) removing magnetic noise from outside the head and compensating for head movements using Temporal Signal Space Separation (tSSS) (Franzmeier et al., 2017); (3) removing SQUID jumps and muscular artifacts through ocular inspection; (4) removing the contribution of ocular and cardiac activity to the MEG signal through the use of the Second Order Blind Identification (SOBI) algorithm (de Oliveira et al., 2021) based on Independent Component Analysis (ICA); (5) segmenting the continuous data into 4-second epochs with 2-second padding on each side to avoid edge effects.

Following data preprocessing, we used a source model to reconstruct the activities of distinct brain locations, referred to as sources. Our source model consisted of a regular grid of 4560 points spaced 1 cm apart forming a cube. 2459 of those were placed inside brain tissue, as specified the Montreal Neurological Institute (MNI) template. For all further analyses, we considered only the 1210 sources belonging to a cortical region according to the Automated Anatomical Labelling (AAL) (Tzourio-Mazoyer et al., 2002).

We linearly transformed the structural information of the source model to the individual 3D image of the head obtained using the Fastrak Polhemus, adjusting the size of the inner skull surface. This allowed us to construct a single shell model, which we used to calculate the lead field matrix through a modified spherical solution (Nolte, 2003).

MEG data were band-pass filtered between 2-45 Hz using a 450^th^-order finite input response (FIR) filter designed with a Hann window. To avoid phase distortion, a two-pass filtering approach was used. In addition, to mitigate edge effects, 2000 samples of real data padding were added to each side of the signal.

Finally, the activity of each participant on each source was reconstructed using a Linearly-Constrained-Minimum-Variance (LCMV) beamformer (Van Veen et al., 1997). This is a widely used technique that spatially filters the MEG data to identify the activity of specific brain sources, while reducing interference from other sources.

### 2.4 Graph construction

A graph is a mathematical representation of a network of elements, called nodes, that are connected by links, known as edges. It is typically expressed as an ordered pair, *G =* (*V, E*), where *V* is a set of nonempty nodes and *E* is a set of edges, each denoting a connection between two nodes. Additionally, edges can have values assigned to them, known as weights, which represent the distance between the nodes an edge connects.

In the context of brain networks, nodes correspond to brain regions, and edges correspond to some sort of connection between them. In this study, nodes correspond to cortical sources from the AAL in five frequency bands independently, and edges correspond to the functional connectivity values between them. Consequently, we ended up using graphs containing 6050 nodes (1210 cortical sources across five frequency bands) and weighted, undirected edges. Despite the added computational complexity, we chose to use sources instead of brain regions and to keep the weights instead of binarizing the graphs to avoid the significant loss of information that these would entail.

We calculated functional connectivity between all nodes using amplitude envelope correlation (Figure 1). This involved four steps: (1) dividing each source signal into five frequency bands (delta: 2-4 Hz, theta: 4-8 Hz, alpha: 8-12 Hz, beta: 12-30 Hz, and gamma: 30-45 Hz), (2) orthogonalizing each source time series to eliminate source leakage, (3) calculating the envelopes of each source activity in each frequency band using the Hilbert transformation (Brookes et al., 2012), and (4) quantifying functional connectivity as the Pearson correlation coefficient between envelopes (Brookes et al., 2011). The absolute values of the coefficients were used as the weights of the edges in the graphs. We refrained from setting a cutoff value for these weights to prevent introducing biases in graph structure that arise from selecting a specific threshold.

**FIGURE 1.**
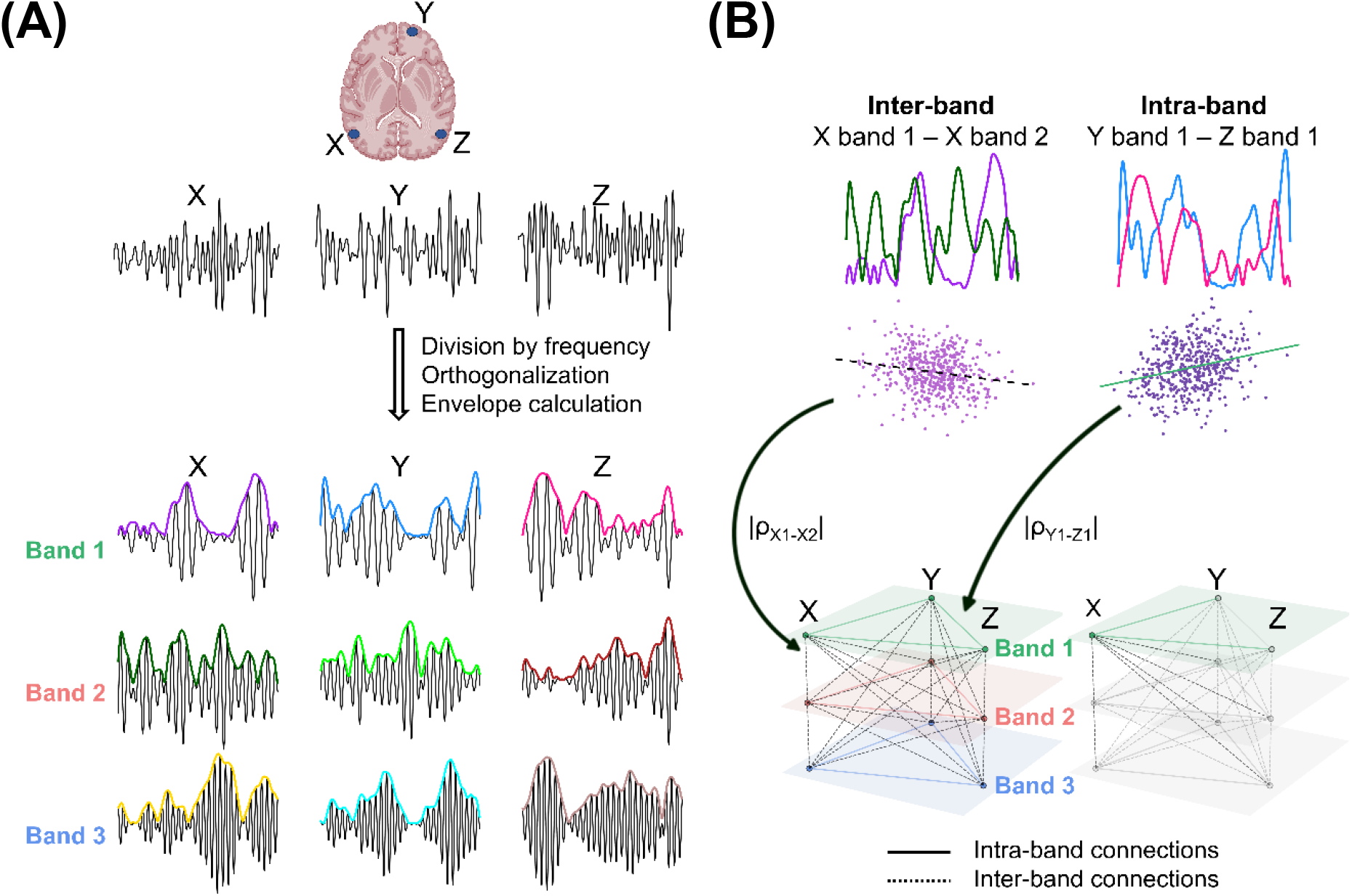
Schema of cross-frequency graphs. For simplicity, we display only three brain sources (X, Y, Z) and three frequency bands (Band 1, Band 2, Band 3), although in the study 1210 brain sources and five frequency bands were used. (A): we calculated the MEG signal of each source, which was then divided by frequency; the signals were then orthogonalized, and the envelopes calculated. The absolute value of the Pearson correlation between envelopes was used as weight for the graph edges. (B): the upper panels show two examples of the correlation calculation: one for inter-band connections and one for intra-band connections; the lower panels display the resulting graph (left) and outline how each node has intra-band connections (solid-colored lines) and inter-band connections (dotted-black lines), accounting for the interplays between bands (right). Although they are represented differently in the figure, both types of edges were treated equally in the centrality metrics’ calculations.

As a result, the brain activity for each participant was represented as a single cross-frequency graph, in which each node was connected to every other node across all frequency bands. This approach allows us to consider both intra- and inter-band edges, as opposed to the more typically employed frequency-specific approaches that ignore inter-band interactions (Figure 1B, lower part).

### 2.5 Centrality calculation

We used three graph measures as centrality indicators: node strength, eigenvector centrality and betweenness centrality. All these measures were calculated for each node of each subject using the NetworkX library (version 2.6.3) and SciPy library (version 1.7.1) in Python (version 3.10.4).

The strength of a node quantifies the node’s direct connections to other nodes in the graph. It is calculated as the sum of the weights of the links incident to node *i*:

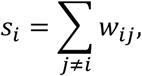

where *w*_*ij*_ is the weight of the link between nodes *i* and *j*.

The eigenvector centrality of a node estimates the relevance of the nodes to which it is directly connected. It is calculated from the following equation:

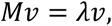

where *M* is the adjacency matrix, *λ* the eigenvalues and *ν* the eigenvectors. The element *i* of the eigenvector *ν* that is associated with the largest eigenvalue *λ*_1_ holds the value for the eigenvector centrality of node *i*.

Finally, the betweenness centrality of a node measures the extent to which the node lies on the shortest paths between other nodes. The shortest path between two nodes is the path with the minimum sum of weights. The betweenness centrality of a node *i* is calculated as the sum of the fraction of the number of shortest paths between all node pairs that pass through node *i*:

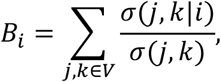

where *V* is the set of nodes, *σ*(*j, k*) is the number of shortest paths, and *σ*(*j, k*|*i*) is the number of those paths passing through some node *i* other than *j, k*. As betweenness centrality takes link weights as a distance measure, we used the inverse of the weights (which, in our case, indicate functional proximity).

To reduce computation time, betweenness centrality was estimated using the algorithm developed by Brandes and Pich (2007). This algorithm allows for the efficient estimation of betweenness centrality by considering only a specified number of random nodes (*k*). In this study, the value of *k* was set to 250, which provided stable estimates.

This resulted in three vectors per subject, each with a length of 6050 (composed of five blocks, each containing 1210 values that correspond to the sources in each frequency band). To obtain a comprehensive measure of centrality, we combined the three metrics into one, hereafter referred to as *centrality score*. To account for the different numeric ranges of the metrics, we first z-scored each of the three vectors independently and then averaged them to obtain a fourth vector of the same length for each subject, the centrality score.

In addition to calculating the centrality score at the frequency band level, we also calculated its band average. To do so, we divided the centrality score vector into its five blocks, each corresponding to a frequency. We then z-scored each block independently, to ensure equal contributions from all bands, and averaged the five scaled blocks, obtaining a single vector of length 1210.

### 2.6 Statistical analysis

The four centrality measures were compared between the control and MCI groups using Matlab (version 2021b). A three-way factorial ANOVA was conducted, with age and years of education as covariables and an alpha level of 0.05. The ANOVA analysis was non-parameterized by using permutations; each F-value obtained for the real data was compared with the F-values obtained for 10.000 randomizations of the original group assignment, which enabled us to calculate the permutation-corrected *p*-value for each F-value.

To account for the large number of comparisons conducted, we employed the two-sided cluster-based permutation test (CBPT), using a Montecarlo approach (Maris and Oostenveld, 2007). Significant sources were grouped in clusters according to their spatial proximity (alpha level of 0.05). Cluster-level statistics were calculated by summing the F-values of all sources within each cluster and comparing that F-value to the null distribution of cluster size, which was obtained through 10.000 randomizations of the group assignments. Only significant clusters after multiple comparisons correction are reported in the results section.

## 3 Results

### 3.1 Classical centrality measures

In this study, we compared strength, eigenvector centrality, and betweenness centrality in each brain source and frequency band between MCI and control participants (Figure 2). It is important to bear in mind that our analyses incorporated cross-frequency interplays to reflect the interactions between oscillatory activity in each source within a given band and all other sources across all bands.

**FIGURE 2.**
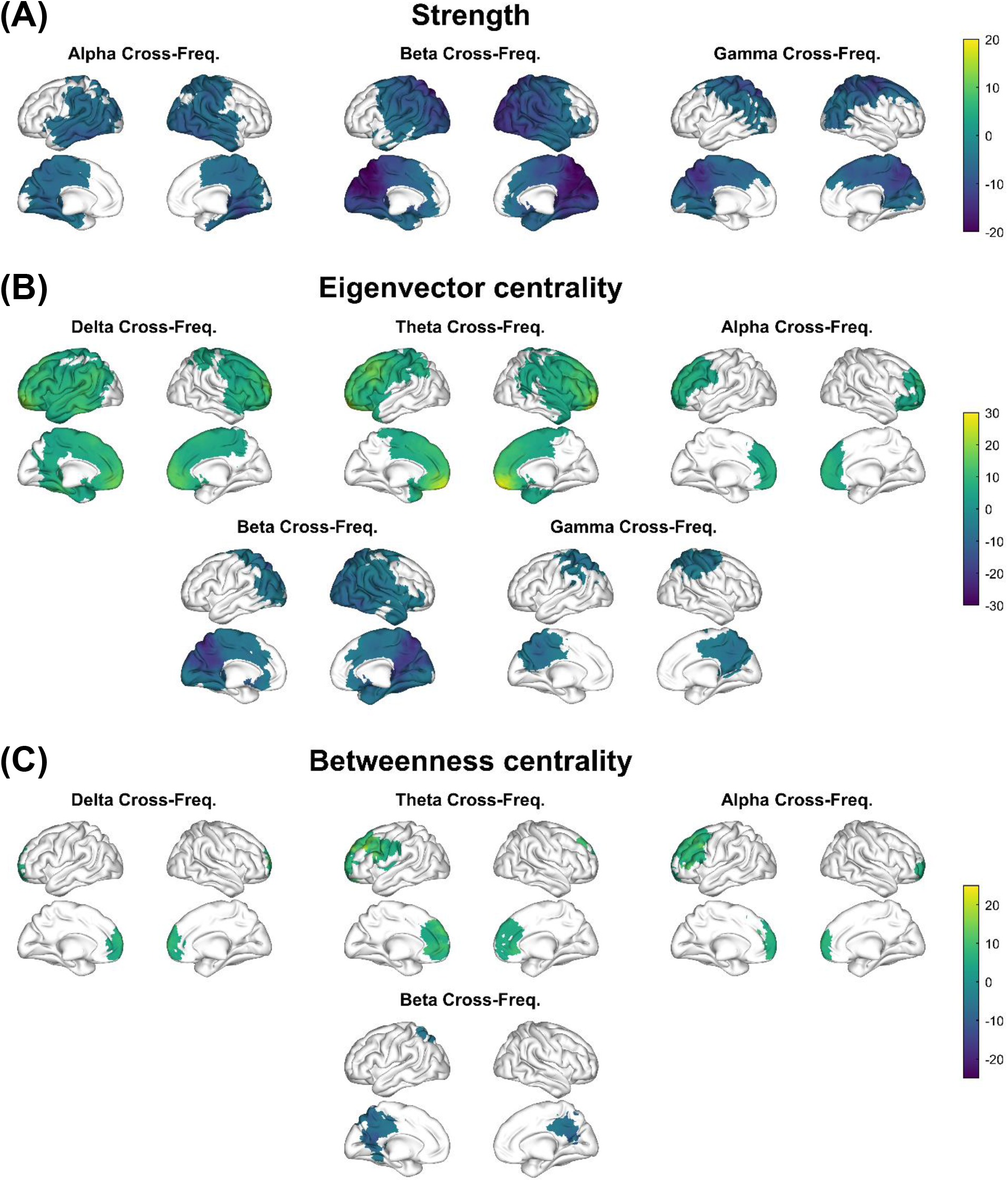
Centrality disruption in MCI assessed with strength (A), eigenvector centrality (B) and betweenness centrality (C). The colored regions indicate significative differences between the MCI and control groups, and the color scale represents the F-value of each source: positive values (greenish colors) denote significant sources with increased centrality in MCIs, while negative values (blueish colors) represent significant sources with decreased centrality in MCIs. For each measure, only bands with at least one significant cluster are shown. The centrality metric of each band includes the links of nodes in that band with the nodes in that same and other bands.

Our results indicate that node strength decreased in the MCI group in various frequency ranges, particularly in posterior and temporal brain regions (Figure 2A). In the alpha band, affected areas included regions posterior to the precentral gyrus (lateral and medial aspects of the parietal and occipital lobes) and the temporal lobe (*F*_*sum*_ *=* 3291, *p*_*cluster*_ *=* 0.0276). The beta band cluster comprised these same areas and included motor regions and the anterior cingulate cortex bilaterally (*F*_*sum*_ *=* 8112, *p*_*cluster*_ *=* 0.0054). Finally, the gamma band showed a decrease in centrality in a smaller portion of the cortex, including the posterior, occipital and posterior part of the frontal lobe (*F*_*sum*_ *=* 3856, *p*_*cluster*_ *=* 0.0158). Notably, all posterior areas of the default mode network were affected, including the hippocampus and parahippocampus bilaterally in alpha and beta, and the precuneus and cingulate gyrus bilaterally in all three bands.

Eigenvector centrality showed a combined pattern of centrality disruption in the MCI group. It increased mainly in the anterior regions in the delta, theta, and alpha bands and decreased in the temporal and posterior areas in the beta and gamma bands (Figure 2B). In the delta band, the centrality increase affected the frontal and parietal lobes, as well as the left temporal lobe (*F*_*sum*_ *=* 5776, *p*_*cluster*_ *=* 0.0032). We found a similar cluster in the theta band, including the same regions except the temporal lobe (*F*_*sum*_ *=* 5545, *p*_*cluster*_ *=* 0.0009). In contrast, in the alpha band, we only found an increase in eigenvector centrality in the frontal lobe (*F*_*sum*_ *=* 1122, *p*_*cluster*_ *=* 0.0064). Concerning decreases in eigenvector centrality, the beta band showed changes in the parietal lobe (extending more anteriorly over the lateral surface of the right hemisphere, including the temporal lobe) and the occipital lobe bilaterally (*F*_*sum*_ *=* 4819, *p*_*cluster*_ *=* 0.0020). Finally, the eigenvector centrality of gamma band showed alterations in a smaller cluster, affecting the precuneus and superior parietal cortex bilaterally (*F*_*sum*_ *=* 1235, *p*_*cluster*_ *=* 0.0304). Once again, the areas of the default mode network were affected: eigenvector centrality increases occurred in the prefrontal and anterior cingulate cortices, while decreases affected the middle and posterior cingulate cortex, precuneus, parahippocampus and right hippocampus.

Finally, betweenness centrality showed a similar dual pattern of increases and decreases in the MCI group, although the clusters were remarkably smaller than in previous metrics. We found a betweenness increase affecting the prefrontal lobe in delta (*F*_*sum*_ *=* 151, *p*_*cluster*_ *=* 0.0102), theta (*F*_*sum*_ *=* 594, *p*_*cluster*_ *=* 0.0002), and alpha (two significant clusters were found: 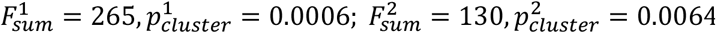), and a decrease over posterior areas of the brain (including the cingulate gyrus and the precuneus bilaterally) in beta (*F*_*sum*_ *=* 208, *p*_*cluster*_ *=* 0.0002).

### 3.2 Centrality score

We integrated the results of the three classical measures into a single measure, the centrality score, ensuring an equal contribution of all bands and metrics. The results include changes in centrality for each band and for the band average (Figure 3).

**FIGURE 3.**
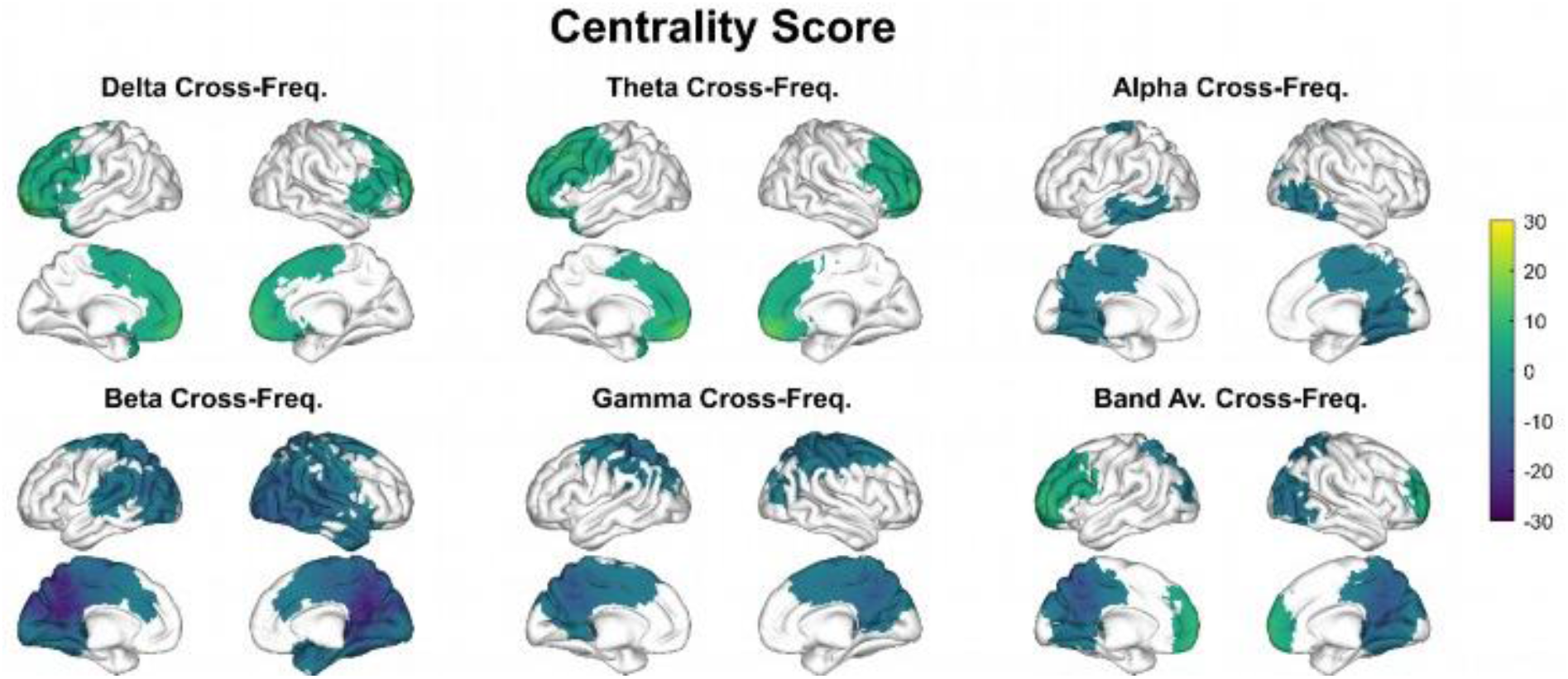
Centrality disruption in MCI assessed with the centrality score. The color code is the same as in Figure 2.

Analysis of individual bands revealed that the MCI group exhibited an increase in centrality in the delta (*F*_*sum*_ *=* 1759, *p*_*cluster*_ *=* 0.0346) and theta (*F*_*sum*_ *=* 2118, *p*_*cluster*_ *=* 0.0126) bands over frontal areas, whereas we observed a decrease in centrality in posterior brain regions across the alpha (*F*_*sum*_ *=* 815, *p*_*cluster*_ *=* 0.0436), beta (*F*_*sum*_ *=* 5542, *p*_*cluster*_ *=* 0.0004) and gamma (*F*_*sum*_ *=* 2238, *p*_*cluster*_ *=* 0.0092) bands. Specifically, centrality reductions affected the parietal lobe in all three bands, the temporal lobe in alpha and beta, and the occipital lobe in beta.

When considering the band average, we observed a combination of the previously observed dual pattern of centrality disruption, with increased centrality affecting the frontal lobe (*F*_*sum*_ *=* 797, *p*_*cluster*_ *=* 0.0284) and decreased centrality affecting temporo-parietal areas (*F*_*sum*_ *=* 1699, *p*_*cluster*_ *=* 0.0090). Remarkably, all the areas of the default mode network were implicated in these results.

## 4 Discussion

This study shows that changes in brain communication patterns in amnestic MCI impact the overall structure of brain networks in two distinct ways: posterior areas reduce their relative importance by decreasing the synchrony between their own high-frequency oscillations and the activity of other regions in all frequency bands, whereas anterior regions increase their importance by synchronizing their low-frequency oscillations with the activity of other regions in all frequencies. Notably, this is the first MEG study to examine network impairment in amnestic MCI using a multilayer, source-based framework.

Electrophysiological studies investigating information flow in the brain typically use a simplified framework, limiting communication analysis to brain regions oscillating within the same frequency bands (Miraglia et al., 2022). However, previous literature has revealed robust interactions between brain regions with different oscillatory rhythms; for example, theta-gamma coupling plays a major role in both working memory (Chuderski, 2016) and long-term spatial memory (Vivekananda et al., 2021). In this sense, our work extends the current understanding of changes in information flow in the early stages of AD by emphasizing the impact of the observed alterations on both intra- and inter-band communication between sources.

Remarkably, and in contrast to most of the previous literature (Miraglia et al., 2022), we constructed source-based graphs that were completely connected and weighted. Despite the technical challenges associated with these large-scale graphs, this holistic approach allowed us to capture the intricate network structure without oversimplification. On these graphs, we assessed centrality changes in MCI. To better understand how information flow changes in the quite complex patterns of brain communication, we created an integrative centrality score that combines three of the metrics most employed in isolation to address this matter (Sporns, 2018), merging the complementary aspects of centrality in the results of our centrality score.

Our analyses comparing the centrality scores between amnestic MCI and control participants reveal two distinct patterns: a spatial pattern and a frequency pattern. The spatial pattern shows that posterior areas reduce their relative contribution to network communication in favor of anterior regions, that show increases in centrality. The frequency pattern shows that the MCI group has decreased centrality through fast-frequency communication and increased centrality through slow-frequency communication.

The result for the band average cross-frequency coupling in the global centrality score metric depicts the above-mentioned spatial pattern, with the MCI group showing a centrality decrease over posterior regions and an increase in anterior areas. This underscores that certain brain regions’ centrality is significantly and consistently compromised in the early course of the disease, regardless of the specific metric or frequency range. These findings align with existing observations on functional connectivity disruptions in MCI due to AD, that report hypoconnectivity in posterior areas and hyperconnectivity in anterior regions (Damoiseaux et al., 2012; Cuesta et al., 2015; López-Sanz et al., 2017; Ranasinghe et al., 2020). Furthermore, the centrality changes we report affect all the main areas of the default mode network, which is consistent with previous studies (Desgranges et al., 2011; Agosta et al., 2012; Jones et al., 2016) showing that these brain regions are particularly vulnerable to AD pathology. However, our results broaden previous evidence suggesting that alterations in functional connectivity translate into changes in the way information propagates through the brain, affecting the overall relevance of extensive brain regions to network communication, as measured by centrality.

The decline in centrality in MCI over posterior areas observed in our results is highly consistent with AD, which entails a disconnection condition (Delbeuck et al., 2003). The abnormal expression of tau primarily contributes to this phenomenon (Ranasinghe et al., 2020), as it affects the cytoskeleton of neurons and leads to synaptic loss (Delbeuck et al., 2003; Ranasinghe et al., 2020). Other studies link hypometabolism with reduced functional connectivity (Smailovic et al., 2020), further supporting these findings.

While disconnection signs have been unanimously interpreted as a pathological sign by previous literature, the biological implications of the network centrality increase we observe in the MCI group over anterior brain areas is subject to more debate. Traditionally, neuroscientists have suggested that functional connectivity increases may palliate some of the burden produced by decreased connectivity in posterior areas of the brain, acting as a compensatory mechanism (Zhang et al., 2010). However, recent research provides an alternative pathological explanation for this phenomenon (Maestú et al., 2021) This hypothesis proposes that Aβ plaques affect preferentially inhibitory neurons, which leads to an increase in excitability, which could result in functional connectivity upregulations. Several studies reinforce this idea by demonstrating that pyramidal neurons in contact with plaques lose GABAergic synapsis (Garcia-Marin et al., 2009), that Aβ causes neuronal hyperactivity (Busche and Hyman, 2020), and that Aβ colocalizes with hypersynchrony, but not with hyposynchrony (Ranasinghe et al., 2020).

Thus, according to this framework, the centrality increase would be interpreted as a pathological sign, contributing to the cascading network failure by increasing neural noise and leading to neuronal death through heightened excitotoxicity. Numerous studies support this theory by showing a widespread loss of functional connectivity among advanced AD patients, including frontal regions (Stam et al., 2009; Wang et al., 2014; Hata et al., 2016). Furthermore, Damoiseaux et al. (2012) performed a longitudinal study in which they measured the functional connectivity of AD patients twice (two to four years apart) using fMRI: the first scan revealed connectivity increases in anterior areas of the default mode network and connectivity decreases in posterior areas, while the second scan showed connectivity decreases in all areas. These findings reinforce hyperexcitability as a precursor to hypoexcitability.

On the other side, the frequency pattern displays centrality increases in low-frequency bands (delta and theta) and centrality decreases in high-frequency bands (alpha, beta, and gamma). It is important to note that the centrality changes we have presented involve the interactions of nodes of one frequency band with each other and with nodes of other bands, as explained in the previous sections. These results suggest that certain brain regions (more concretely anterior areas) increase their relevance in the network by increasing the coupling of its own slow wave activity with other parts of the network, regardless of the latter’s frequency range. Nonetheless, our results are congruent with previous findings on functional connectivity in MCI and AD, that link increments in synchrony mainly to delta and theta and decrements to alpha and beta activity (Núnez et al., 2019; Ruiz-Gómez et al., 2019; Briels et al., 2020; Ranasinghe et al., 2020; Smailovic et al., 2020). In addition, some studies report hyperconnectivity in the alpha range over anterior regions (López-Sanz et al., 2017), whereas hypoconnectivity has rarely been associated with gamma (Cuesta et al., 2022).

Interpreting the specific meaning of the frequency pattern emerging from our results is challenging due to the limited understanding of the human resting state dynamics and the role different frequency bands play in AD. Previous studies have found an increase in power for the delta and theta bands, as well as a decrease for the alpha and beta bands (Huang et al., 2000; López et al., 2014; Cuesta et al., 2015). This pattern may relate to a loss of synapses, which can impede communication between brain regions, as information can flow through fewer “pathways”. In this regard, Cabral et al. (2022) used a computational model to demonstrate how reductions in coupling strength can lead to increased slow-frequency activity. In line with this hypothesis, several studies have reported decreased activity of both theta (Wang et al., 2014) and even delta (Hata et al., 2016) during later stages of the disease progression. In this vein, our results could reflect an intermediate state in which some regions exhibit a diminished ability to contribute to network communication while oscillating in faster rhythms, whereas others show an increased relevance in slower frequencies, resembling the pattern of alterations suggested by the mentioned studies.

When analyzing each individual metric, we observe that both the spatial and frequency patterns are apparent. This was expected, as the three metrics represent centrality. Nevertheless, it is worth noting that the metrics reflect different aspects of a node’s significance: node strength measures the number and intensity of connections a node has, eigenvector centrality considers the relevance of said connections, and betweenness centrality estimates the capacity of a node to connect other nodes in the network through shortest paths.

As a result of these nuances, each metric yields unique results. The results for node strength only show decreases in alpha, beta, and gamma bands. Remarkably, most of the brain is affected, even the posterior parts of the frontal lobe. This might be due to the neuronal death that AD entails. Remarkably, no increases in strength were observed, meaning that none of the regions exhibit a significant increase in the magnitude of its raw total connection to the rest of the network.

The results for eigenvector and betweenness centrality show increases in delta, theta, and alpha, alongside decreases in beta (both metrics) and gamma (only eigenvector centrality). These findings reinforce the loss of coordination in the posterior brain regions, but also highlight heightened centralization within the frontal cortex. This might be due to the disconnection occurring among posterior areas, which appears responsible for shifting relative importance towards anterior brain regions – a phenomenon previously reported (Huang et al., 2000; Engels et al., 2015).

Another noteworthy feature is the variable centrality changes displayed by the alpha band, depending on the metric. This finding might suggest a transitional frequency role between faster and slower brain rhythms for the alpha band in the maintenance of the global communication structure. While the role of the beta and theta activity is consistently altered towards decreased and increased centrality respectively, alpha band activity seems to display both patterns depending on the aspect of centrality we pay attention to.

Lastly, it is remarkable that betweenness centrality alterations in the MCI group affect a smaller portion of the network than changes in the other metrics. This is likely due to betweenness centrality being a more global measure that considers all other nodes in the network –reflecting higher-scale aspects of centrality– while strength and eigenvector centrality are more local, depending mainly on a node’s immediate neighbors. Thus, the smaller disruption in betweenness centrality may indicate that while local changes are pronounced, the overall information flow structure of the network is less affected. This observation underscores the importance of considering different centrality measures to gain a comprehensive understanding of global network dynamics.

The main limitation of this study is the lack of biomarkers, leading to some uncertainty about the presence of AD in all subjects with MCI. However, it is worth noting that all patients exhibited memory impairment, which is strongly linked to the subsequent development of AD in particular (Dubois and Albert, 2004; Schmidtke and Hermeneit, 2008). However, future studies should include biomarker data to better characterize the relationship between centrality changes and disease progression in AD.

Overall, our findings shed light on the complex changes that occur in brain networks in the early stages of AD. The results indicate a noteworthy shift in centrality distribution from posterior to anterior regions, indicating a decline in relative significance of the former and an increase in importance of the latter. These findings demonstrate a remarkable level of consistency, with congruent outcomes across all bands and metrics, suggesting that using a unified approach that considers the interactions between different frequency bands can enhance our understanding of AD network dynamics in a more comprehensive manner. Furthermore, this study establishes a new approach to comprehensively study disruptions in cross-frequency functional networks not only for AD, but also for other neurological disorders studied through electrophysiology techniques.

## Data Availability

All data produced in the present study are available upon reasonable request to the authors

## Acknowledgements

This study was supported by EU Horizon 2022 framework program funding to ‘eBRAIN-Health’ project (https://www.ebrains.eu/) to Ignacio Taguas.

